# Age- and sex-related changes in motor functions: A Comprehensive Assessment and Component Analysis

**DOI:** 10.1101/2024.04.04.24305334

**Authors:** Veronika Wunderle, Taylan D. Kuzu, Caroline Tscherpel, Gereon R. Fink, Christian Grefkes, Peter H. Weiss

## Abstract

Age-related motor impairments often cause caregiver dependency or even hospitalization. However, comprehensive investigations of the different motor abilities and the changes thereof across the adult lifespan remain sparse.

We, therefore, extensively assessed essential basic and complex motor functions in 444 healthy adults covering a wide age range (range 21 to 88 years). Basic motor functions, here defined as simple isolated single or repetitive movements in one direction, were assessed by means of maximum grip strength (GS) and maximum finger-tapping frequency (FTF). Complex motor functions, comprising composite sequential movements involving both proximal and distal joints/muscle groups, were evaluated with the Action Research Arm Test (ARAT), the Jebsen-Taylor Hand Function Test (JTT), and the Purdue Pegboard Test.

Men achieved higher scores than women concerning GS and FTF, whereas women stacked more pins per time than men during the Purdue Pegboard Test. There was no significant sex effect regarding JTT. We observed a significant but task-specific reduction of basic and complex motor performance scores across the adult lifespan. Linear regression analyses significantly predicted the participants’ ages based on motor performance scores (R^2^ = .502). Of note, the ratio between the left- and right-hand performance remained stable across ages for all tests. Principal Component Analysis (PCA) revealed three *motor components* across all tests that represented dexterity, force, and speed. These components were consistently present in young (21 – 40 years), middle-aged (41 – 60 years), and older (61 – 88 years) adults, as well as in women and men. Based on the three *motor components*, K-means clustering analysis differentiated high- and low-performing participants across the adult life span.

The rich motor data set of 444 healthy participants revealed age- and sex-dependent changes in essential basic and complex motor functions. Notably, the comprehensive assessment allowed for generating robust *motor components* across the adult lifespan. Our data may serve as a reference for future studies of healthy subjects and patients with motor deficits. Moreover, these findings emphasize the importance of comprehensively assessing different motor functions, including dexterity, force, and speed, to characterize human motor abilities and their age-related decline.

## Introduction

Aging is a physiological process that leads to a heterogeneous but progressive decrease in motor functions. The age-related decrease in motor functions often increases dependency, morbidity, and even hospitalization (Gielen et al., 2023; Proietti & Cesari, 2020). Consequently, there is an increasing demand to characterize healthy motor aging (Keevil & Romero-Ortuno, 2015; World Health Assembly, 2020). In this context, Frangos and colleagues stressed the importance of assessing multimodal functionality (Frangos et al., 2023).

Upper limb function, especially hand function, is a crucial marker for a healthy and independent life (Incel et al., 2009). Previous studies described hand function as a promising marker for physical abilities and successful rehabilitation, cognition, general health across the lifespan, and even quality of life (Forrest et al., 2018). Notably, the established assessments to quantify upper limb function have been mainly validated by comparing healthy individuals to patients with motor deficits (Kwakkel et al., 2019). Grip strength and dexterity are predominantly used to characterize motor deficits in patients with neurological disorders such as Parkinson’s disease or stroke (Hensel et al., 2019; Scherbaum et al., 2020). In contrast, studies investigating comprehensively the physiological decline of motor functions during healthy aging, including assessing diverse basic and complex abilities, remain sparse. A particular challenge to this endeavor is the variability of motor functions among individuals. Furthermore, there are sex-specific differences for specific motor performance scores. For example, on average, men outperform women regarding grip strength, while women exhibit superior finger dexterity (Dodds et al., 2014; Gómez-Campos et al., 2022; Vasylenko et al., 2018).

Despite the growing demand, only a limited number of studies have assessed upper limb motor functions multi-dimensionally, considering the various functional demands that the upper limb needs to address in daily life (Kobayashi-Cuya et al., 2018). Furthermore, the relationships among different motor abilities often remain unexplored (McKay et al., 2016, 2017b). Moreover, limited sample sizes and restricted age distributions make evaluating specific age- or sex-related effects difficult. Bravell and colleagues, for example, comprehensively analyzed motor function in a population older than 60 years and found that, especially in women, the decline of motor functions including fine motor abilities, balance/strength and flexibility were associated with mortality (Bravell et al., 2017). Rueckriegel and colleagues focused their assessment on a population younger than 18 years and on writing and drawing tasks. They found that repetitive movements matured earlier than more complex fine motor function (Rueckriegel et al., 2008). Both studies highlighted the importance of a multidimensional approach regarding (hand-) motor function.

Extensive databases, such as the NKI-Rockland Sample, UK-Biobank, or the Rotterdam study, predominantly focus on grip strength or pegboard-test performance. Thus, due to the limited amount of applied motor tests, a detailed analysis of the different factors contributing to upper limb motor function is not feasible in these cohorts (Nooner et al., 2012; Sudlow et al., 2015; van der Willik et al., 2020). Bowden and McNulty also emphasized the need for multidimensional assessments. They measured grip strength, grooved-pegboard performance, and finger-tapping speed in a cohort of 70 healthy participants (20 - 88 years, 35 females). Besides the expected age-dependent changes in these three motor tests, the authors showed that performance in these motor tests varied independently across age and sex. However, they did not analyze the relationship between the different motor tests (Bowden & McNulty, 2013).

Therefore, the current study aimed to comprehensively assess upper limb motor function in a relatively large cohort of healthy participants, considering age and sex as key factors. We employed a battery of well-established motor tasks addressing different aspects of hand motor function, which have been validated in healthy individuals and patient cohorts. The test battery comprised basic motor tasks such as grip strength and finger tapping speed, as well as complex motor tasks such as the Purdue Pegboard Test, the Jebsen-Taylor Hand Function Test, and the Action Research Arm Test (ARAT) (Allgöwer & Hermsdörfer, 2017; Aoki & Fukuoka, 2010; Desrosiers et al., 1995b; Rehme et al., 2011; Tscherpel et al., 2019). Besides analyzing age- and sex-related effects on basic and complex motor functions, we investigated the relationships between individual performance scores of the different motor tests by applying principal component analysis (PCA). Finally, a K-means clustering approach was adopted to detect deviations from the normal healthy aging-associated decrease in motor performance. This approach enabled the categorization of high- and low-performing healthy individuals across the adult life span.

## Methods

### 1. Participants

A total of 474 adults were recruited by the Motor Assessment Center of the Collaborative Research Center 1451 (Fink et al., 2022) of the German Research Foundation (DFG) from April 2021 to October 2023. The following inclusion criteria were applied for the current study: age between 21 and 90 years, non-left-handedness (defined as a laterality quotient larger than −28 as determined by the Edinburgh Handedness Inventory; Oldfield, 1971), the absence of any relevant depressive symptoms (operationalized by unremarkable results in the Beck Depression Inventory (BDI-II, ≤ 13), (Kühner et al., 2007) and the Montgomery-Åsberg Depression Rating Scale (MADRS ≤ 12) (Montgomery & Asberg, 1979)), as well as having no prior history of neurological, psychiatric, or orthopedic diseases (selection of participants in Figure 1). The final sample consisted of 444 healthy participants (mean age of 52.5 years ± 18.5 years, with a range of 21 to 88 years; 254 [57.2%] females) (descriptive statistics in supplementary Table S1). Trained examiners administered a standardized motor ability assessment in two laboratories at either the Department of Neurology of the University Hospital of Cologne or the Institute of Neuroscience and Medicine (INM-3 – Cognitive Neuroscience) at the Research Center Jülich. All participants provided written informed consent. The ethics committee of the Medical Faculty of the University of Cologne approved the study performed under the Declaration of Helsinki.

**Figure 1:**
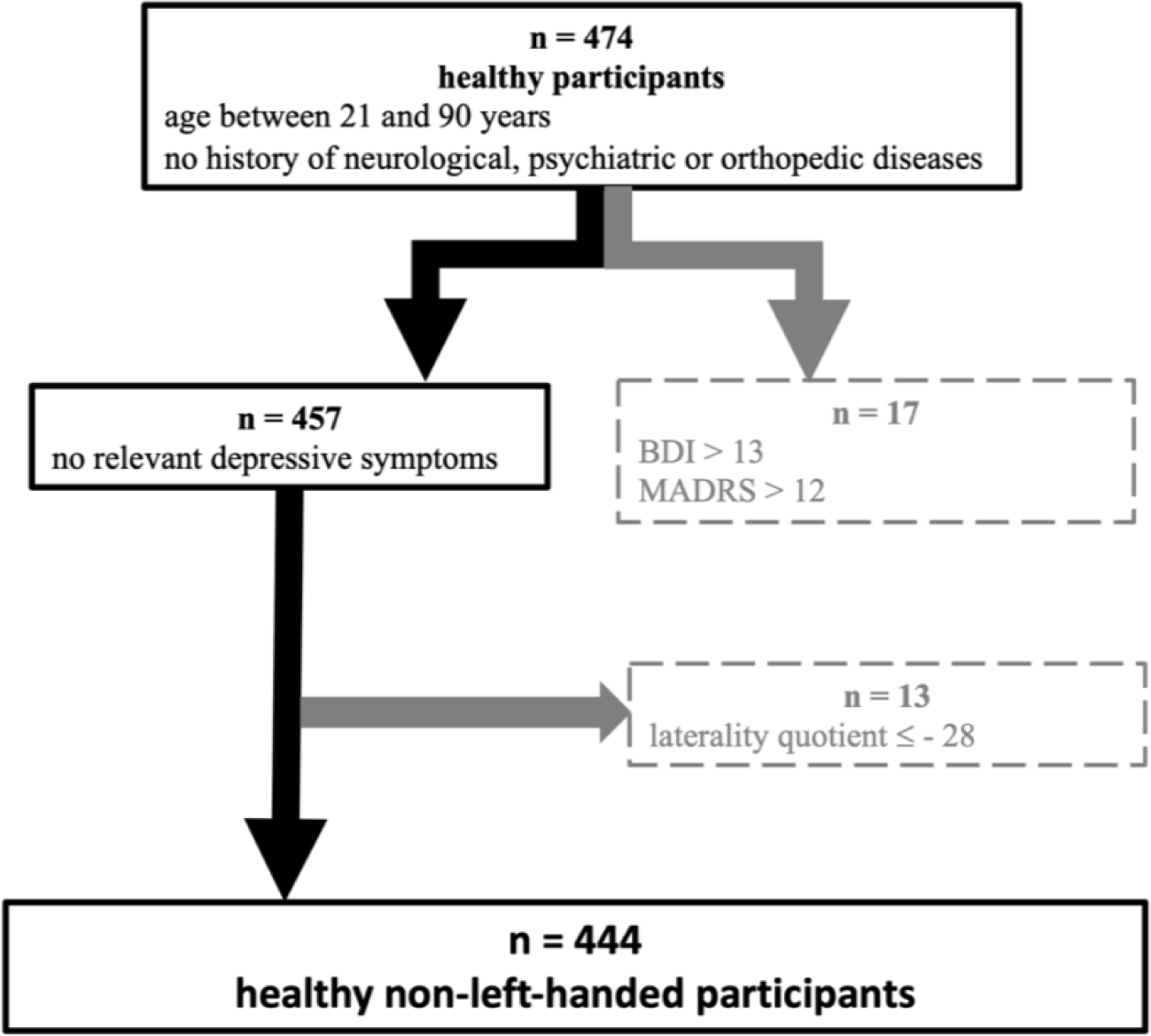
Flowchart depicting the selection of participants. From 474 participants, who were screened for eligibility, we finally included 444 individuals as healthy, non-left-handed participants. Exclusion criteria are depicted in the grey boxes on the right side.

### 2. Motor Assessment

To account for the broad age range and interindividual variability in motor performance, we employed five robust and reliable motor tests to assess individual motor abilities comprehensively (Kwakkel et al., 2019) and a measure of handedness. All tests were administered by three trained study nurses and according to a standardized test protocol including detailed written instructions to be read out to the participants to guarantee comparability. Additionally, there were weekly meetings to discuss occurring questions or uncertainties regarding test administration or interpretation.

#### 2.1 Edinburgh Handedness Inventory

The participant‘s handedness was determined using the Edinburgh Handedness Inventory (Oldfield, 1971). Based on the preferred hand for ten activities of daily living, a laterality quotient (LQ) was calculated. Participants with an LQ score greater than – 28 were included in the subsequent analyses.

#### 2.2 Action Research Arm Test (ARAT)

The ARAT is commonly used to evaluate upper limb functions and related disabilities in neurological patients. It consists of four subtasks, i.e., grasp, grip, pinch, and gross movement, reliably accounting for the different aspects of upper limb function (ICC = 0.98). (Hsieh et al., 1998; Lyle, 1981; Nomikos et al., 2018).

#### 2.3 Maximum Grip strength

Maximum grip strength was assessed using a vigorimeter (KLS Martin Group, Germany), which measures the force applied on a rubber ball grasped by the participant with a whole-hand grip (Rehme et al., 2011; Volz et al., 2016).

This highly standardized method measures the force of combined finger flexion and hand closure, minimizing compensatory movements (ICC = 0.92) (Desrosiers et al., 1995a; Sipers et al., 2016). Three trials were performed for each hand in alternating order to avoid fatigue. The mean of the three trials for each hand was computed for further analysis. The relative grip strength was calculated (grip strength non-dominant hand / grip strength dominant hand) to account for interindividual differences in absolute strength.

#### 2.4 Purdue Pegboard Test

The Purdue Pegboard Test is a reliable and valid method for evaluating arm and hand function and finger dexterity (ICC = 0.81 - 0.89)(Buddenberg & Davis, 2000; Reddon et al., 1988). Participants were instructed to insert as many pins as possible into designated holes on a rectangular board within 30 seconds. Three trials were performed with each hand in alternating order, and the average of these three trials was utilized for further analysis (Desrosiers et al., 1995b). Additionally, the relative Purdue Pegboard Test performance was calculated (numbers of pins non-dominant hand / number of pins dominant hand) to adjust for interindividual differences.

#### 2.5 Jebsen-Taylor Hand Function Test (JTT)

The JTT is a reliable tool for evaluating upper limb function through simulated daily living activities such as eating or drinking (ICC = 0.84 – 0.97) (Jebsen et al., 1969; Sığırtmaç & Öksüz, 2020). Six standardized one-handed subtasks were administered, including turning over cards, picking up small objects, simulated feeding, stacking checkers, lifting large light objects, and lifting large heavy objects. Similar to other studies, we excluded the task “writing”, as it depends on motor function and cognitive/educational abilities (Diekhoff-Krebs et al., 2017; Hummel et al., 2005). Participants were instructed to complete each task with their dominant and non-dominant hand as quickly as possible. The time taken to complete each subtask was recorded and summed up for each hand separately. Additionally, the relative JTT performance was calculated (total time non-dominant hand / total time dominant hand) to adjust for interindividual differences in the absolute time needed to complete the tasks.

#### 2.6 Maximum Finger-Tapping Frequency (FTF)

The FTF task (Wang et al., 2009) was designed to measure isolated fast repetitive movements of the index finger. Participants were instructed to repeatedly press a button with their index finger as quickly as possible. An arrow on a computer screen indicated the hand to be used for tapping, and a dice with a height of 2.5 cm indicated the required tapping amplitude. Each trial lasted 2 s and was followed by a slightly jittered, on average 3 s pause to avoid fatigue. The finger-tapping frequency was calculated separately for each hand as the average of three trials (Tscherpel et al., 2019, 2020). The relative finger-tapping frequency was calculated (finger-tapping frequency non-dominant hand / finger-tapping frequency dominant hand) to account for interindividual differences in absolute speed.

### 3. Statistical analysis

Statistical analyses were performed using IBM SPSS Statistics (Statistical Package for the Social Sciences, version 29, SPSS Inc., Chicago, Illinois, USA) and Prism (GraphPad Software, LLC, Version 9.4.1). For all analyses, a significance level of p < 0.05 was set. If not otherwise stated, data are depicted as mean ± standard error of the mean (SEM).

#### 3.1 Correlation analysis

Pearson correlation analyses were performed and Bonferroni-corrected for multiple comparisons to test for relationships between motor performance and age.

#### 3.2 Linear regression analysis

We used a linear regression analysis to predict the participant’s age based on motor performance. Accordingly, age was used as the dependent variable. For each hand separately, the following variables were entered simultaneously as the eight independent (predictor) variables and checked for collinearity: maximal grip strength [kPa], maximal finger-tapping frequency [Hz], number of stacked pins in the Purdue Pegboard Test, and total time needed for the Jebsen-Taylor Hand Function Test [s] (Smith, 2018).

#### 3.3 Group comparisons

To evaluate the effects of age and sex on motor performance, we performed a repeated measures analysis of variance (ANOVA) with the between-subject factors ‘AGE’ and ‘SEX’ and the within-subject factor ‘HAND’ on the performance for the following tests with the dominant or non-dominant hand: grip strength, maximum finger-tapping frequency, Purdue Pegboard Test, and Jebsen-Taylor Hand Function Test. The between-subject factor SEX had two levels: women and men. For the between-subject factor AGE, the whole sample of 444 participants was divided into three age groups: 153 participants (88 women, 57.5%) with ages between 21 and 40 years constituted the young group, 106 participant (72 women, 67.9%) with ages between 41 and 60 years were considered as middle-aged, and 185 participants with ages between 61 and 88 years comprised the older group (94 women, 50.8%) (Voelcker-Rehage, 2008).

Two-sided t-tests were used *post-hoc* to further characterize significant effects. These t-tests were Bonferroni-corrected for multiple comparisons. Besides, we report the effect sizes. We conducted a *post-hoc* power analysis to confirm a sufficient power of our calculations (G*Power version 3.1, (Faul et al., 2007)4/4/2024 3:27:00 PM. Performing repeated measures ANOVA with a sample size of n = 444, we achieved a power of 99% for detecting at least medium effects at a significance criterion of α = .05.

#### 3.4 Principal component analysis (PCA)

All test scores were z-transformed and subjected to a principal component analysis (PCA) to evaluate the correlational pattern between the performances in the different motor tests. The Kaiser-Meyer-Olkin (KMO) measure of sampling adequacy (Kaiser, 1974) and Bartlett’s test of sphericity (Bartlett, 1951) indicated that the current data set fulfilled the requirements for PCA. Only components with eigenvalues > 1 were extracted, per the Kaiser criterion. The varimax rotated component matrix was used to examine each test’s relative contributions (i.e., components loadings) on the extracted components. Loadings ≥ 0.4 were considered reliable, and ≥ 0.7 were regarded as meaningful for a given component (if not cross-loading onto another component) (C. C. Schmidt et al., 2022). The resulting principal components were considered to reflect different motor dimensions required for the task set loaded on a given component. Therefore, the components were termed *motor components*.

Based on the younger group of 153 participants (i.e., 21 to 40 years old), we calculated weighted component scores for each of the three motor components using regression. The resulting individual component scores represented the participants‘ relative position along the three motor components (meaning their weighted performance in the tests that cluster on the respective component (Halai et al., 2017)). The individual weighted component scores were used as behavioral variables for separating two subgroups within the whole sample (n= 444) using K-means clustering.

## Results

### 1. Overview of motor performance

To provide an overview of the motor performance for the entire sample of the 444 non-left-handed healthy participants, we report the results for the basic and complex motor tests as well as the ratios between the non-dominant hand’s performance and dominant hand’s performance collapsed across age groups and sexes in Table 1.

**Table 1:** Overview of motor test performance (n = 444) Listed are the mean values and standard deviations (SD) of the motor test performances of the whole sample (n = 444).

| Motor test performance | mean | SD |
| --- | --- | --- |
| Purdue Pegboard Test (dominant hand) | 14.10 pins | 2.28 |
| Purdue Pegboard Test (non-dominant hand) | 13.24 pins | 2.19 |
| Jebsen-Taylor Hand Function Test (dominant hand) | 27.84 s | 4.72 |
| Jebsen-Taylor Hand Function Test (non-dominant hand) | 29.40 s | 5.26 |
| Grip strength (dominant hand) | 61.51 kPa | 17.02 |
| Grip strength (non-dominant hand) | 58.59 kPa | 17.48 |
| Finger-tapping frequency (dominant hand) | 9.54 Hz | 1.14 |
| Finger-tapping frequency (non-dominant hand) | 9.55 Hz | 1.05 |
| Ratios between non-dominant and dominant hands |  |  |
| Purdue Pegboard Test-Ratio | 94.25% | 9.04 |
| Jebsen-Taylor Hand Function Test-Ratio | 106.35% | 10.47 |
| Grip strength-Ratio | 95.34% | 11.00 |
| Finger-tapping frequency-Ratio | 93.82% | 11.71 |

With respect to the tests probing more complex motor functions, the ARAT scores showed no variance. All 444 participants achieved the maximal total score of 57 points. Hence, ARAT was excluded from all further analyses, given the strong ceiling effects of this test in healthy participants, which did not add additional information to the assessment of motor performance.

### 2. Reduced motor performance in aging

Pearson correlation analysis was applied for the parameter of a given motor test and age as a continuous variable to assess the motor performance in the different motor tests across the adult life span. There was a significant decline in performance in all basic and complex motor tasks across the examined age range (21 - 88 years) (Figure 2). For clarity, we only report the results of correlation analyses for the dominant hand in the text. The results of the correlation analyses for the non-dominant hand are documented in Table 2. Grip strength (p < .001, r = -.349) and FTF (p < .001, r = -.369) demonstrated a significant negative correlation with age. In addition, the number of stacked pins in the Purdue Pegboard Test decreased significantly with age (p < .001, r = -.641). As expected, the time required to perform the JTT exhibited a significant positive correlation with age (p < .001, r = .491).

**Figure 2:**
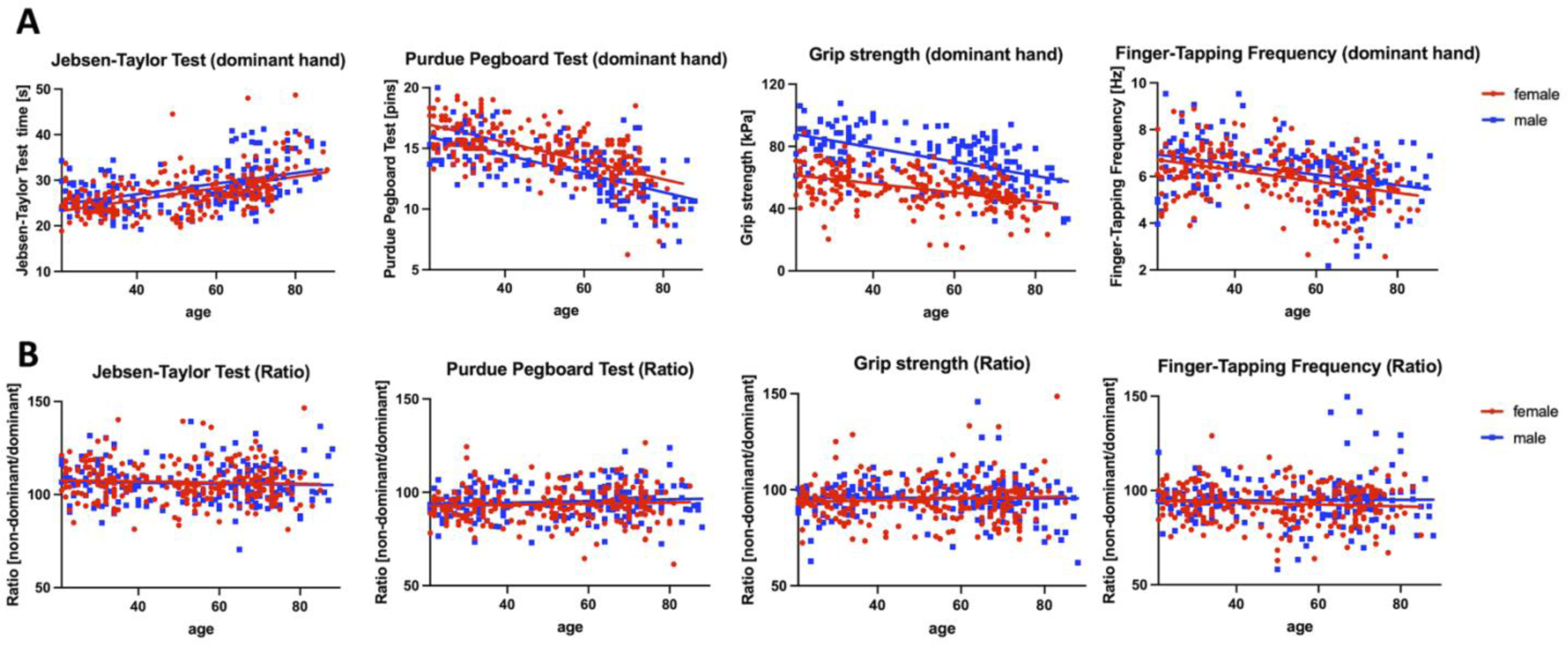
Depiction of the correlation of motor tasks and age and the age-independent performance ratios. **(A)** Significant correlation between age and the respective motor scores (Jebsen-Taylor Test, Purdue Pegbaord Test, grip strength, and finger-tapping frequency) performed with the dominant hand. Female participants (red) and male participants (blue) are shown separately. (**B)** Age-independent performance ratios (performance of non-dominant hand divided by performance of dominant hand) are depicted. Female participants (red) and male participants (blue) are shown separately.

**Table 2:** Correlation analyses between age and motor test performance and ratios. Results of the Bonferroni-corrected Pearson correlation analysis revealed a significant correlation between the age and the performances of the 444 healthy participants in the four motor tests performed with their dominant and non-dominant hands. In contrast, there was no significant correlation between age and the ratios between the test performances of the non-dominant and the dominant hands for any of the four motor tests.

| Motor test performance | Pearson correlation coefficient | p-value |
| --- | --- | --- |
| Purdue Pegboard Test (dominant hand) | -.641 | < .001 |
| Purdue Pegboard Test (non-dominant hand) | -.577 | < .001 |
| Jebsen-Taylor Hand Function Test (dominant hand) | .491 | < .001 |
| Jebsen-Taylor Hand Function Test (non-dominant hand) | .438 | < .001 |
| Grip strength (dominant hand) | -.349 | < .001 |
| Grip strength (non-dominant hand) | -.313 | < .001 |
| Finger-tapping frequency (dominant hand) | -.369 | < .001 |
| Finger-tapping frequency (non-dominant hand) | -.415 | < .001 |
| **Ratios between non-dominant and dominant hands** |  |  |
| Purdue Pegboard Test-Ratio | .082 | >.999 |
| Jebsen-Taylor Hand Function Test-Ratio | -.052 | >.999 |
| Grip strength-Ratio | .035 | >.999 |
| Finger-tapping frequency-Ratio | -.033 | >.999 |

Notably, the performance ratios of the four basic and complex motor tests (performance with the non-dominant hand divided by the performance of the dominant hand) did not demonstrate any significant correlation with age (all p > .999) and thus remained stable across the examined age range (21 – 88 years, see Table 2).

### 3. Effects of age and sex on motor performance

For the basic motor tests (grip strength, FTF) and the more complex motor tests (Purdue Pegboard Test, JTT), we calculated repeated measures ANOVAs with the within-subject factor HAND (dominant, non-dominant) and the between-subject factors AGE (young (21-40 years), middle-aged (41 - 60 years), older (61 - 88 years)) and SEX (men, women). We included the detailed statistics and the *post-hoc* tests in Table 3.

**Table 3:**
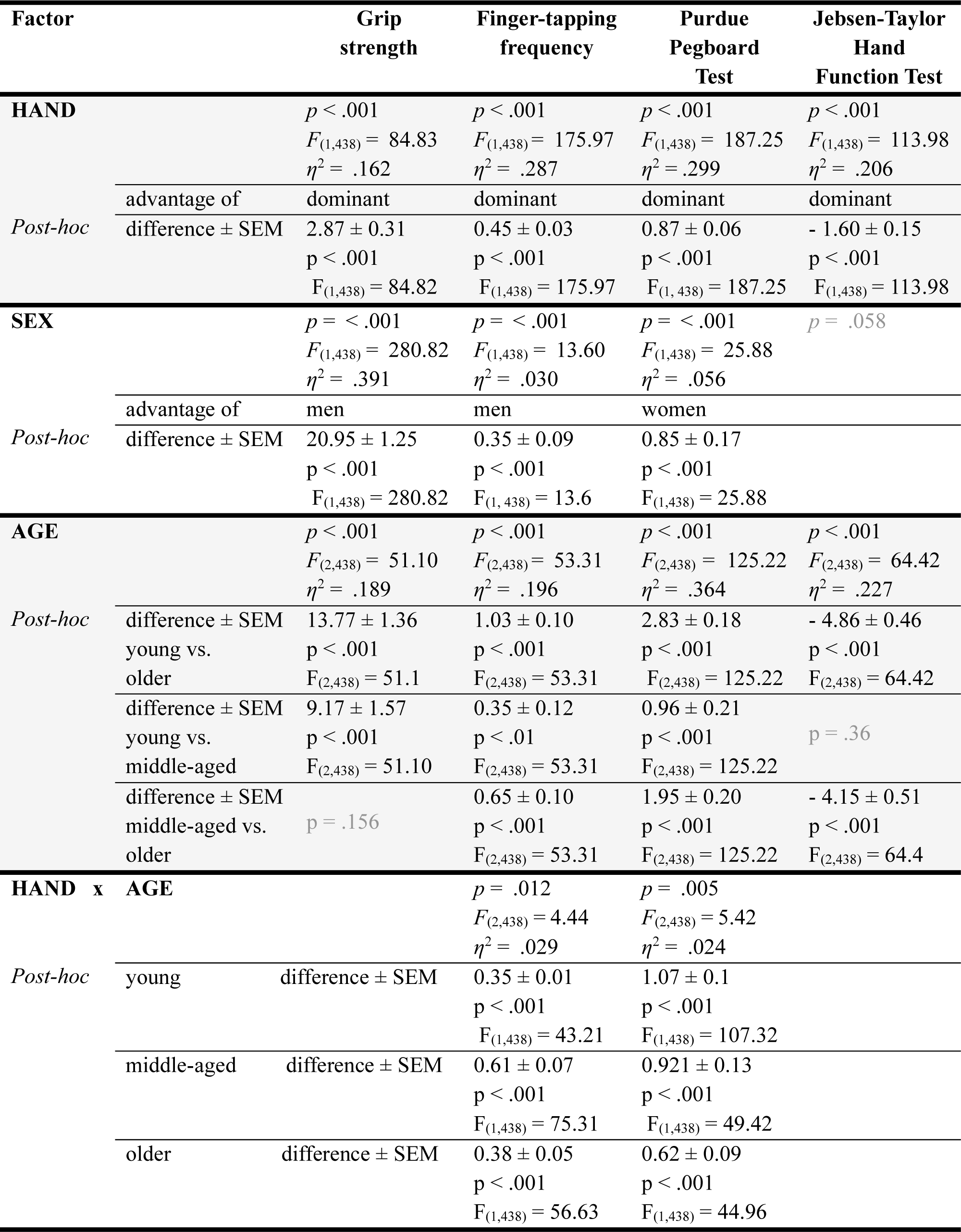
Results of repeated-measures ANOVA. Results of the repeated-measures ANOVA revealed significant effects for the within subject factor HAND and the between-subject factors SEX and AGE. Additionally, the Bonferroni corrected statistics of the *post-hoc* t-tests are depicted.

There were significant main effects for the within subject factor HAND and the between subject factor AGE for the basic motor tests (grip strength, FTF) as well as for the complex motor tests (Purdue Pegboard Test, JTT).

Additionally, we observed main effects for the between subject factor SEX for three motor tests (grip strength, FTF, Purdue Pegboard Test). Notably, the JTT revealed only a trend for the factor SEX.

*Post-hoc* comparisons indicated that the performance of the dominant hand was better than that of the non-dominant hand regarding every test.

Additionally, men achieved higher scores than women concerning grip strength and FTF, whereas women stacked more pins per time than men during the Purdue Pegboard Test. Moreover, young participants outperformed older participants in all scores.

Notably, the pattern of this age-dependent decline differed within the tests. There was a consistent age-dependent reduction regarding Purdue Pegboard Test performance and FTF. Grip force decreased significantly from young to middle-aged participants. However, there was no further significant decrease for older participants (Figure 3).

**Figure 3:**
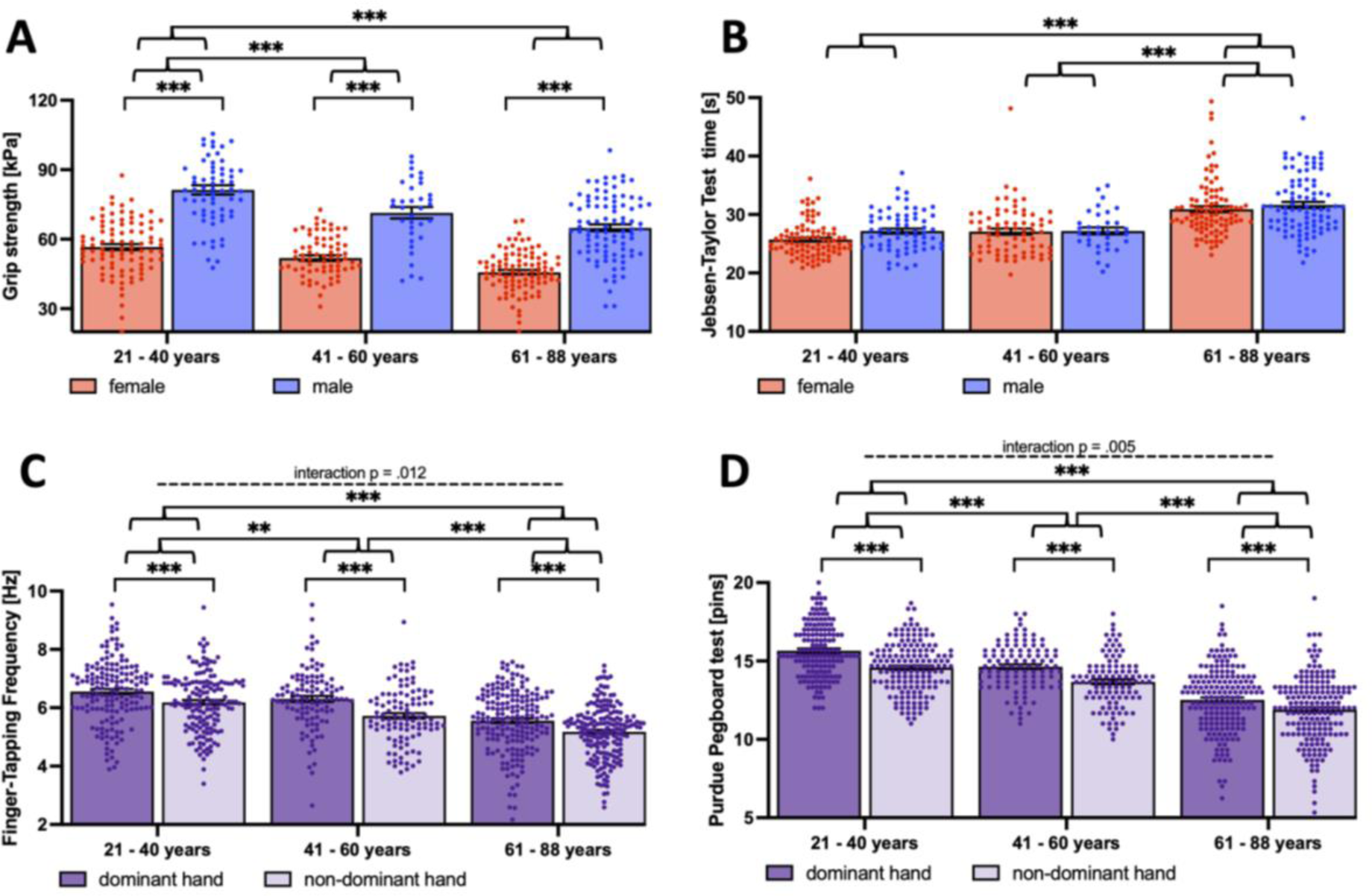
Depiction of the significant AGE (young, middle-aged, older), SEX (women, men), and HAND (dominant, non-dominant hand) effects and interactions. Displayed are the results of the repeated measures ANOVA evaluating AGE, SEX, and HAND effects. **(A)** Significant SEX and AGE effect for grip strength. Depicted are the average grip strengths of both hands for female (red) and male (blue) young, middle-aged, and older participants. **(B)** Significant AGE effect for Jebsen-Taylor Test. Depicted is the average time needed per test of both hands for female (red) and male (blue) young, the middle-aged, and older participants. **(C)** Significant HAND and AGE effect, as well as the interaction of AGE and HAND for finger-tapping frequency. Depicted are the tapping frequencies of the dominant (dark purple) and non-dominant (light purple) hands for young, middle-aged, and older participants. **(D)** Significant HAND and AGE effect, as well as interaction of AGE and HAND for Purdue Pegboard Test. Depicted are the stacked pins of the dominant (dark purple) and non-dominant (light purple) hands for young, middle-aged, and older participants. * = p<0.05, ** = p<0.01, *** = p<0.001, **** = p<0.0001

JTT, on the other hand, showed stable performance scores for the young and middle-aged groups, but a significant decrease from middle-aged to older participants (see Figure 3 and Table 3 for post-hoc tests).

There was a significant interaction of the factors HAND x AGE for the performance in the Purdue Pegboard Test and FTF (see Figure 3 and Table 3 for statistics and post-hoc tests). Regarding the Purdue Pegboard Test, the difference between dominant and non-dominant hands became less pronounced with increasing age.

For FTF, the interaction was driven by a higher difference between dominant and non-dominant hands for the middle-aged.

For the current cohort of 444 healthy participants, we provide descriptive statistics (i.e., mean, median, range, SD, SEM) for the performance in the four motor tests with either the dominant or non-dominant hands in supplementary Table S2. Based on the repeated measures ANOVA, values are given for six groups of participants comprising young (age between 21 and 40 years) as well as middle-aged (age between 41-60 years) and older (age between 61 and 88 years) women and men. These values could be used to compare single-subject data to the respective control group (Crawford et al., 1998).

### 4. Predicting age by motor performance

A multiple linear regression analysis investigated whether motor performance significantly predicted the participants’ age. Grip strength, Purdue Pegboard Test and JTT performances, and FTF were utilized as predictors, while age served as the dependent variable. Indeed, the overall regression model yielded statistically significant results (Adjusted R^2^ = .502, F_(8,435)_ = 56.854, p < .001), indicating that the predictors collectively had a significant effect on the prediction of age. Specifically, grip strength (β = -.290, p = 0.003), performance in the Purdue Pegboard Test (β = -.462, p < .001), and finger-tapping frequency (β= -.166, p = 0.005) emerged as significant predictors of age.

### 5. Motor components derived from PCA

To disclose potential components underlying the performance in the different basic and complex motor tests, we conducted a PCA on the motor scores of both hands for the entire cohort. This approach identified three principal components, which collectively accounted for 86.2% of the total variance observed. Component 1 (termed motor component “dexterity”), explaining 48.1% of the variance, included strong loadings from the Purdue Pegboard Test and the JTT. The grip strength of both hands loaded on component 2 (termed motor component “force”) accounted for 22.3% of the variance. Finally, component 3 (termed motor component “speed”), explaining 15.8% of the variance, was related to the FTF of both hands. Importantly, none of these tests showed reliable loadings on multiple components. Table 4 provides the specific loadings of each test.

**Table 4:** Principal component analysis (PCA) revealing three robust motor components across the adult lifespan (n = 444) Depicted is the rotated component matrix for the three motor components (dexterity, force, speed) resulting from the performances’ principal component analysis (PCA) when the 444 healthy participants performed four motor tests with their dominant and non-dominant hands. The three components accounted together for 86.2% of the total variance. Component loadings (after varimax rotation) above 0.7 are highlighted and printed in bold, with the corresponding (sub)tests considered relevant for that component.

|  | Component<br>1<br>(*dexterity*) | Component<br>2<br>(*force*) | Component<br>3<br>(*speed*) |
| --- | --- | --- | --- |
| Purdue Pegboard Test (dominant) | **0.88** | 0.04 | 0.12 |
| Purdue Pegboard Test (non-dominant) | **0.90** | 0.05 | 0.11 |
| Jebsen-Taylor Test (dominant) | **- 0.86** | - 0.11 | - 0.19 |
| Jebsen-Taylor Test (non-dominant) | **- 0.83** | - 0.12 | - 0.24 |
| Grip strength (dominant) | 0.10 | **0.97** | 0.10 |
| Grip strength (non-dominant) | 0.08 | **0.96** | 0.12 |
| Finger-tapping frequency (dominant) | 0.21 | 0.06 | **0.93** |
| Finger-tapping frequency (non-dominant) | 0.21 | 0.15 | **0.91** |
| Explained variance | 48.1 | 22.3 | 15.8 |
| (after rotation) in % | (38.7) | (24.4) | (23.0) |
| Eigenvalues (after rotation) | 3.85 (3.10) | 1.79 (1.95) | 1.26 (1.84) |

As we have encountered age-dependent differences in performance, we also performed separate PCAs for the age groups (21-40 years, 41-60 years, and 61-88 years). Remarkably, the partition into the three components and their respective assignment remained unchanged (variance explained young: 80.4%, middle-aged: 78.9%, older: 84.8%, see Table S3 for details), proving a robust separation into the three fundamental *motor components* dexterity (JTT and Purdue Pegboard Test), (grip) force, and speed (FTF).

As a next step, we calculated sex-specific PCAs (women and men; see Table S4). The patterns observed for the entire cohort were replicated for both groups. The three fundamental *motor components* dexterity, force, and speed explained 84.7% and 85.4% of the total variance in women and men, respectively.

In summary, the PCA of motor performance parameters exposed a clear separation into three fundamental *motor components*, which were robust across sex and age.

### 6. K-means Clustering

We extracted individual component scores based on the three *motor components* (dexterity, force, speed) identified by the above-described PCA. These scores represent the participant’s relative position along the principal component based on their weighted performance in the respective assessments. The young group, including young women and men, served as the reference. Due to the heterogeneity of motor performance, we aimed to group the participants in an unbiased and data-driven way, using K-means clustering. This approach generated two subgroups of healthy participants without relying on subjective criteria like age or fixed thresholds, appreciating individual strengths and weaknesses of the participants.

The first cluster comprised 148 (33.3%) participants, while the second cluster comprised 296 (66.7%) participants. Within the young group, 13 participants (8.5%) were assigned to the first cluster, whereas 21 (19.8%) of the middle-aged group and 114 (61.6%) of the older group were assigned to the same cluster. 140 young (91.5%), 85 middle-aged (80.2%), and 71 (38.4%) older participants were assigned to the second cluster. Notably, significant differences were observed between these healthy subgroups, indicating a clear separation between high-performing participants (second cluster) and those displaying reduced performance in specific motor tests (first cluster, see Figure 4).

**Figure 4:**
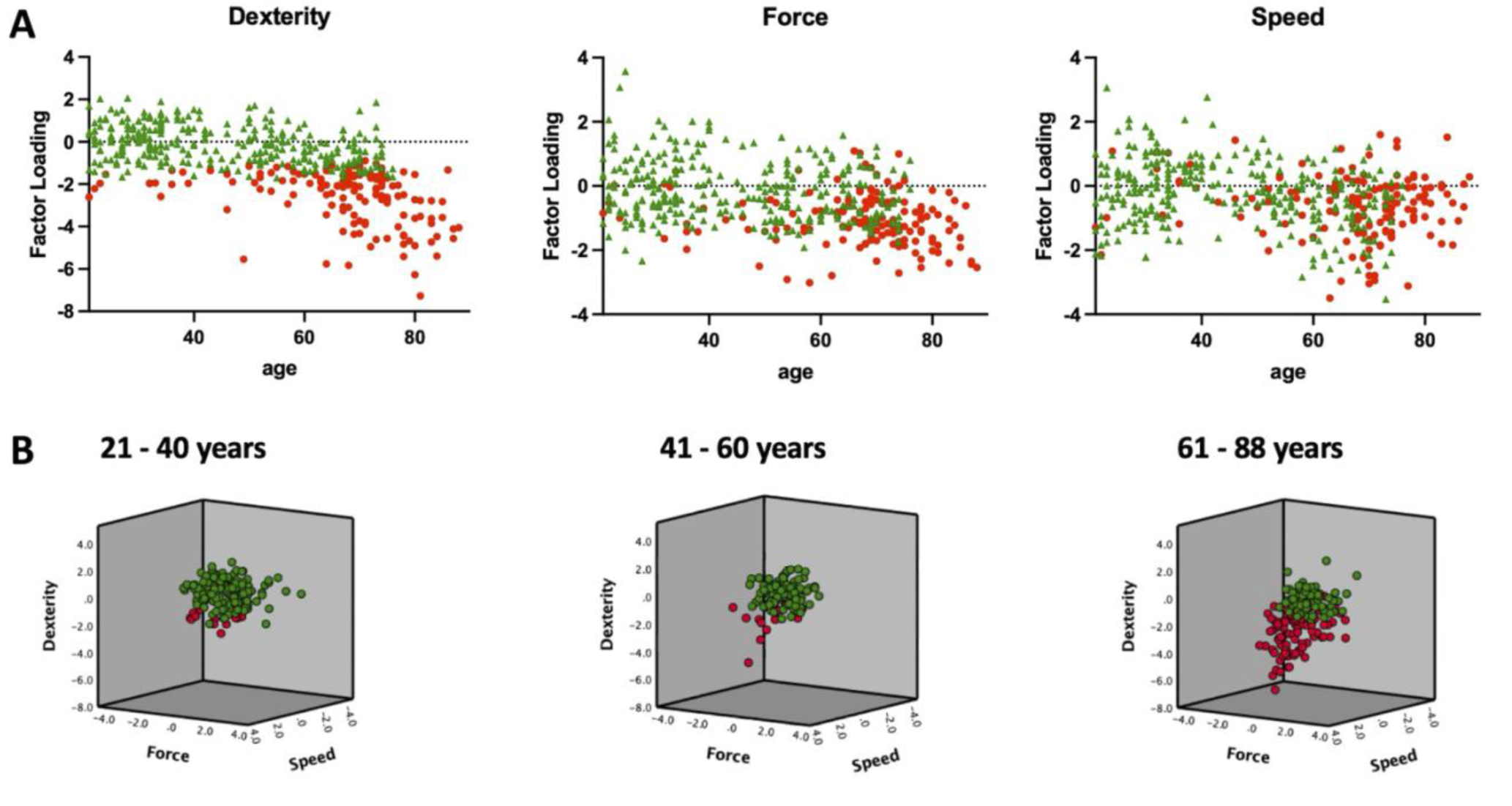
K-means clustering based on the three motor components revealed two groups / clusters of high- and low-performing participants. The results of the data-driven separation of high- and low-performing participants using K-means clustering are displayed. **(A)** Individual factor loadings of all participants’ motor components (dexterity, force, speed) are depicted in relation to their age. Participants of the second cluster perform well in all given components (green, n = 296), while participants of the first cluster show inferior performance in at least one motor component (red, n = 148). **(B)** The three-dimensional graph combining all three motor components illustrates the separation of high (green) and low (red) performing young (21 - 40 years), middle-aged (41 - 60 years), and older participants (61 - 88 years).

## Discussion

A comprehensive motor assessment examining essential basic (grip strength, finger-tapping frequency) and complex (Purdue Pegboard Test, Jebsen-Taylor Hand Function Test, Action Research Arm test) motor functions of the upper limb in a large cohort (n = 444) of healthy participants covering a broad age range (21 – 88 years) revealed an age-dependent reduction in motor functions, along with pronounced sex-specific differences across the lifespan. Notably, the ratios between the performance of the dominant and non-dominant hands within each test remained stable. Using linear regression, motor performance (especially grip strength and Purdue Pegboard Test and FTF) predicted the age of the participants. By applying Principal Component Analysis (PCA), three robust *motor components*, namely dexterity, force, and speed, were identified that remained stable across lifespan for men and women. These three *motor components* served as the basis for a K-means clustering approach, which allowed to distinguish high- and low-performing participants.

The current study assessed basic motor functions via grip strength and finger-tapping frequency. Grip strength is one of the most commonly used measures of overall muscle strength depending on muscle mass and neuromuscular mechanisms. Age-dependent changes in this context include reduced cortical excitability and nerve conduction velocity or the loss of alpha-motorneurons, but also impaired agonist and antagonist activation coordination, resulting in reduced voluntary muscle contraction (Carson, 2018; Clark, 2019). Finger tapping speed was selected since this simple motor test can be applied across a broad age range and is sensitive to age-related decline of motor functions in healthy participants (Hubel et al., 2013; Tscherpel et al., 2019, 2020).

The assessment of complex motor functions comprised the Purdue Pegboard Test, the Jebsen-Taylor Hand Function test, and the Action Research Arm test (ARAT). The longitudinal study by van der Willik and colleagues observed an age-dependent decrease in Purdue Pegboard Test performance starting from 56 years with no sex effect (van der Willik et al., 2020). In contrast, Vasylenko and colleagues observed an age-dependent slowing of movement in the Purdue Pegboard Test, which was more pronounced in men (n = 49) than in women (n = 51) (Vasylenko et al., 2018). Our data replicated the previously described age-related performance reduction in the Purdue Pegboard Test. Notably, this reduction was already observed between the young (21 - 40 years) and the middle-aged (41 – 60 years) participants. We, furthermore, confirmed sex differences in the Purdue Pegboard Test, with women outperforming men. Unlike Vasylenko and colleagues, we did not find age-dependent sex differences. This discrepancy might be due to different readouts, as we counted the number of inserted pins instead of assessing the kinematics involved (Vasylenko et al., 2018). However, there was a significant interaction between the hand used for the pegboard test and age. The difference in pegboard performance between the dominant and non-dominant hands became less pronounced with age.

The Jebsen-Taylor Hand Function Test evaluates various hand functions necessary for daily living activities. Again, hand function decreases with age in men and women (Hackel et al., 1992).

Finally, the ARAT is a well-established, hand and arm function test used in neurological patients. It contains different demands on fine- and gross upper limb motor functions (Alt Murphy et al., 2015; Lyle, 1981; Pohl et al., 2020). Notably, there was a ceiling effect for all healthy participants examined in the current study, since all participants achieved the maximum score of 57 in the ARAT. In turn, this means that deviations from the ARAT’s maximum score are indicative of pathological motor behavior. Thus, the strength of the ARAT rather lies in detecting and monitoring pathological motor performance than assessing complex motor behavior in general.

The application of these diverse tests allowed us to consider multiple motor functions and thus comprehensively characterize motor abilities in a large cohort of healthy women and men (n = 444). This approach sets us apart from studies on extensive databases, focusing on one or very few motor tests (Nooner et al., 2012; Sudlow et al., 2015). Therefore, we additionally offer normative values for all performed motor tests regarding different age groups and both sexes in a German-Speaking population. To our knowledge, there is no comparable data available yet.

### 1. Reduction of motor performance in aging

The study replicated the age-related reduction in grip strength, finger tapping speed, and performances in the Purdue Pegboard and the Jebsen-Taylor Hand Function tests. These findings align with prior studies investigating these motor tests separately (Aoki & Fukuoka, 2010; Desrosiers et al., 1995b; Dodds et al., 2014; van der Willik et al., 2020).

Given that aging is a complex process, many factors contribute to a general decline in motor functions. They include the physiological reduction in muscle mass (Soyuer et al., 2023), reduced coordination or neuromuscular impairments (Carson, 2018; Clark, 2019), as well as changes in sensory, visual, and attention capabilities (Heintz Walters et al., 2021).

Importantly, we could show differential, task-specific patterns of motor performance decline with age (see Figure 2).

Both the FTF and the Purdue Pegboard test revealed a consistent performance decline for each age group, which aligns with prior studies (Hubel et al., 2013; Stijic et al., 2023). In contrast, the reduction of grip force was only significant between young (21 - 40 years) and middle-aged (41 - 60 years) participants. There was no significant grip force difference between middle-aged and older participants. This finding is in common with Dodds and colleagues, as well as Gómez-Campos and colleagues, who described a peak grip force around the age of 40 years and a following decline (Dodds et al., 2014; Gómez-Campos et al., 2022). Nevertheless, another pattern appeared for the JTT, for which the performance was stable in the groups of young and middle-aged participants. The JTT performance reduction became significant when comparing the middle-aged and older groups. A potential reason for this pattern might be the resemblance of the JTT subtasks with activities of daily living (ADL), which are limited in older participants only.

### 2. Age-independent asymmetry ratios

Remarkably, our investigation revealed that the performance ratios between the non-dominant and dominant hands did not exhibit any relevant age-dependent changes. Across all age groups, the dominant hand consistently outperformed the non-dominant hand. This finding aligns with the findings of Cosby and colleagues, who obtained comparable grip strength ratios in 214 participants with an age range of 16 years to 63 years (Crosby et al., 1994). Additionally, we could show that the hand performance ratios were similar across the different tests, indicating a high reliability of the hand performance ratios as a potential robust metric of proper hand function.

Although performance asymmetries, to a certain extent, do not need to indicate abnormalities (Pool et al., 2014), increased asymmetry indices of hand function can potentially serve as an early indicator of evolving motor deficits. For instance, Parker and colleagues associated hand-grip strength asymmetry of more than 20% with an increased likelihood of future limitations in instrumental activities of daily living (IADL) in 18235 Americans older than 50 years (Parker et al., 2021). This finding is further supported by research linking asymmetries of motor functions with functional limitations (Collins et al., 2020), morbidity (Klawitter et al., 2022), risk of future falls (McGrath et al., 2021), and even cognitive deficits (Jia et al., 2023).

Moreover, we observed a significant interaction of HAND (dominant versus non-dominant hand) and AGE for the Purdue Pegboard Test performance. At generally lower performance levels, older participants (61-88 years) showed less pronounced differences between their dominant and non-dominant hand performances compared to middle-aged (41-60 years) and young participants (21-40 years) (Figure 3). These findings confirm but also extend (in a different age range) previous results in a sample of 98 right-handed participants ranging from 5 to 24 years (Roy et al., 2003). These authors also showed “a right-hand advantage in performance which was larger in the younger than the older participants” in the Annett pegboard task.

Notably, there was an additional interaction of HAND and AGE regarding FTF. While young and older participants showed similar differences between the finger-tapping frequencies of the dominant and non-dominant hands, this difference was more pronounced in the middle-aged group. This pattern could be potentially explained by a differential age-related decrease in the finger-tapping performance of the two hands. If the decline of finger-tapping frequency of the non-dominant hand starts earlier (already in the middle-aged group) than that of the dominant hand (present in the older group), then the difference in finger tapping between the non-dominant and dominant hands would be maximal in the middle-aged group.

Thus, the previous and our current findings underscore the importance of examining absolute hand performance scores and considering the relative performance between the dominant and non-dominant hands as valuable research and potential clinical parameters.

### 3. Prediction of age by motor performance

Regarding the robust correlation between motor performance and age, we conducted a linear regression analysis, utilizing the motor scores to predict age. Grip strength, Purdue Pegboard Test performance, and FTF significantly predicted age, underlining the close interplay of different motor abilities. Consistent with our results, Martin and colleagues described an increasing association between strength and dexterity, which gets more pronounced with age (Martin et al., 2015). Pérez-Parra and colleagues performed multiple linear regression analyses associating grip strength with the JTT, the Nine-Hole Peg test, and aging (Pérez-Parra et al., 2023). Notably, their findings align with our PCA results, as every *motor component* (dexterity, force, and speed) was significantly represented in our regression analysis, emphasizing the importance of a multidimensional approach to assess motor function.

With an R^2^-value of 0.502, our linear regression model still contains relevant unexplained variance. One reason might be the age-dependent increasing variability caused by “high-and low-performing” participants’ as described in the section on K-means clustering. That performance variability increases with age has been shown for finger-tapping frequency (Hubel et al., 2013). Such an age-dependent variability-increase challenges a linear regression model, leading to more unexplained variance, i.e., reducing the predictive power of the linear regressions.

### 4. Sex effects on motor performance

The current comprehensive multidimensional assessment of motor function uncovered sex-dependent variations in performance across the different motor tests. Specifically, men exhibited significantly higher grip strengths and finger-tapping-frequencies, while women performed significantly better in the Purdue Pegboard Test. This pattern of results aligns with the 1000 Norms project, which suggested that men performed better in gross motor tasks while females outperformed men in dexterous tasks (McKay et al., 2017b, 2017a). Similarly, studies claimed that men are faster in repeating a single movement, while women outperform men in the speed of programming a sequence of hand movements (Nicholson & Kimura, 1996; S. L. Schmidt et al., 2000). It has been suggested that these performance patterns may be associated with testosterone-mediated fast-twitch muscle fibers or morphological asymmetry patterns in the brain, which were mainly described for the cerebellum (Fan et al., 2010; Hubel et al., 2013; Shigehara et al., 2022), an issue that warrants further investigation.

Interestingly, there was no significant sex effect regarding the JTT. Different strategies can be used to accomplish the JTT subtasks, as suggested by kinematic analysis during JTT performance (Kontson et al., 2020). Thus, men and women might use different strategies that, nevertheless, result in similar JTT performance.

### 5. Robust motor components across the adult lifespan differentiate low- and high-performer

Principal component analysis (PCA) revealed three *motor components* – namely dexterity, force, and speed - persisting throughout the entire lifespan and regardless of sex. Notably, the basic motor assessments grip strength and FTF separated into two independent *motor components*, while the complex tasks Purdue Pegboard test and JTT formed one joint *motor component*. The separation of the different motor tasks was data-driven and did not depend on artificial classifications of motor assessments.

The three robust *motor components* underline the importance of considering different motor functions when assessing motor performance, as they prove the existence of different and independent motor abilities. In this vein, Bowden and McNulty already showed, that strength, speed, and dexterity need to be evaluated independently (Bowden & McNulty, 2013). Yet, they did not show whether the motor tasks represented different and independent motor functions.

In line with this, Ingram and colleagues introduced a physiological profile assessment (PPA), a multimodal test battery to assess independent motor functions (Ingram et al., 2019). It measured grip strength, reaction time, dexterity, tactile sensation, bimanual coordination, and a functional task. Putting these tests in relation to each other, the authors described four domains of upper limb function: gross motor skills, arm stability, fine motor control, and tactile discrimination (Ingram et al., 2023). Our study confirms and extends these previous findings by demonstrating that the separation of *motor components* remains independent of age and sex. This allows for an analysis of motor function across the whole population through the comparison of *motor components*.

In contrast to Ingram and colleagues, who calculated a composite score from the average motor performances to discriminate between healthy and impaired participants, we used a data-driven K-means-clustering approach. This approach was independent of previously built hypothesis or cut-off values. K-means clustering based on the three *motor components* revealed those participants in the cohort whose motor performance was disproportionally low. Within each age group, it successfully segregated individuals into two distinct subgroups with overall low (first group) or high (second group) performance levels in the applied tests of motor functions. Thus, the importance of a multidimensional assessment of motor function is also highlighted by the results of the K-means clustering approach. For example, participants who performed well in force or speed could still be assigned to the “low-performing” group due to substantially reduced dexterity. The current approach enables the discrimination between orthogonal agers, who maintain their motor abilities in advanced age, and disproportional agers, who show a pronounced reduction in motor function. Importantly, this approach does not define pathological behavior. However, recognizing a disproportionate reduction in upper limb motor function (i.e., over and above the reduction that is expected by that person’s age) could serve as an early indicator of abnormal aging. Therefore, K-means clustering by motor performance might help to detect age-related syndromes, such as frailty or sarcopenia, which are associated with an increased risk of help-dependency and morbidity (Umegaki, 2016).

The concept of ‘Ageing well’ has been declared a global health priority by the World Health Organization, drawing attention to late-life health (Keevil & Romero-Ortuno, 2015). Identifying low-performing individuals with (even mild) motor impairments offers the opportunity to provide specific support and interventions, potentially preventing further decline in motor abilities (Talar et al., 2021). A longitudinal design would allow observing whether the low-performing participants are at specific risk for diseases/morbidity. The current data allows identifying ‘healthy’ individuals with reduced motor functions based on the (normative) values provided for a large cohort of participants (n=444) with a broad age range (21-88 years; see Table S2). Moreover, the current data facilitate the detection of deficiencies in patients with neurological (or other) diseases across different motor dimensions by using the documented stable ratios between the dominant and non-dominant hands or the ARAT.

## Conclusion

This study provides a comprehensive assessment of age- and sex-dependent motor functions in a large cohort of healthy participants. It highlights the added value of multidimensional approaches and puts established motor scores in relation to each other, generating fundamental *motor components,* which can help to identify disproportional decline in motor function.

## Limitations

As we included only German-speaking non-left-handers (LQ > −28) of the German population, the current results are not generalizable to left-handed individuals. Moreover, the reported normative values are specific for the examined population of only German-speaking non-left-handers and cannot directly be generalized to other populations in Europe or elsewhere.

The current study design is cross-sectional, without a follow-up measurement regarding the longitudinal development of the individual participant. Further studies with longitudinal designs are warranted to track the individual development of motor functions. However, due to organizational issues, such longitudinal studies will always be hampered by a restricted time interval for follow-ups.

We aimed for an efficient motor assessment in a relatively large cohort of participants covering a broad age range. Thus, we did not acquire kinematic data during the performance of the test battery. Kinematic analyses might have brought additional insights into healthy movement patterns (Hensel et al., 2022), but the time-consuming kinematic recordings would have significantly reduced the size of the tested cohort Notably, the ARAT did show a pronounced ceiling effect, as all healthy participants obtained the full score of 57 points. Thus, the ARAT seems to provide an important measure to rather discriminate between healthy and pathological states of the motor systems. It is used as reliable measure in studies working with different patient groups, including stroke, traumatic brain injury, cerebellar ataxia or multiple sclerosis (Pike et al., 2018; Platz et al., 2005; Reoli et al., 2021; Saini et al., 2021). On the other hand, the ARAT does not seem to yield meaningful results in a healthy population.

Focusing on the comprehensive assessment of hand function, we did not include measures of other (i.e., lower limb) motor functions such as locomotion or balance. Additionally, integrating cognitive tasks might bring additional value to investigating the motor system’s healthy aging. Thus, future studies may consider a combined assessment of lower and upper limb functions, including dual-task paradigms (e.g., performing a cognitive task in parallel with a motor task).

Finally, we would like to clarify that the use of k-means clustering to separate high- and low performing participants does not intent to identify pathological behavior. However, it can help to detect disproportional agers who might suffer from frailty or sarcopenia and thus might profit from specific support to maintain health. Longitudinal studies are needed in order to characterize further the long-term impact of the classification by k-means clustering of motor performance.

## Acknowledgements

Funded by the German Research Foundation (DFG) - Project-ID 431549029. We thank Ulrike Freiburg, Sabrina Lentfort, Susanne Ossig, and Natascha Kellner for technical support.

## Data availability

All data produced in the present study are available upon reasonable request to the authors.

