## Supplementary Tables for "Age- and sex-related changes in motor functions: A Comprehensive Assessment and Component Analysis"

**Table S1.**

**Descriptive statistics of the cohort (n = 444).**

| | Group [sex (age)] | n | mean | SD | SEM | min. | max. |
|---|---|---|---|---|---|---|---|
| **Age** | female (21–40) | 88 | 30.08 | 5.08 | 0.54 | 21 | 40 |
| | male (21–40) | 65 | 30.61 | 5.59 | 0.69 | 21 | 40 |
| | female (41–60) | 72 | 51.99 | 5.29 | 0.63 | 41 | 60 |
| | male (41–60) | 34 | 52.59 | 5.85 | 1.00 | 41 | 60 |
| | female (61–88) | 94 | 70.91 | 5.68 | 0.59 | 61 | 88 |
| | male (61–88) | 91 | 70.98 | 6.60 | 0.70 | 61 | 87 |
| | all (21–88) | 444 | 52.50 | 18.50 | 0.88 | 21 | 88 |
| **laterality - quotient (LQ) of the EHI** | female (21–40) | 88 | 82.17 | 19.92 | 2.11 | 0 | 100 |
| | male (21–40) | 65 | 80.08 | 21.44 | 2.64 | -20 | 100 |
| | female (41–60) | 72 | 84.00 | 17.13 | 2.03 | 0 | 100 |
| | male (41–60) | 34 | 78.88 | 21.93 | 3.77 | 30 | 100 |
| | female (61–88) | 94 | 88.19 | 17.38 | 1.79 | 10 | 100 |
| | male (61–88) | 91 | 84.13 | 25.37 | 2.67 | -10 | 100 |
| | all (21–88) | 444 | 83.57 | 20.73 | 0.98 | -20 | 100 |
| **BDI-II** | female (21–40) | 88 | 3.06 | 3.30 | 0.35 | 0 | 12 |
| | male (21–40) | 65 | 3.08 | 3.22 | 0.40 | 0 | 13 |
| | female (41–60) | 72 | 2.80 | 3.52 | 0.42 | 0 | 13 |
| | male (41–60) | 34 | 3.03 | 3.16 | 0.54 | 0 | 11 |
| | female (61–88) | 94 | 3.64 | 3.42 | 0.35 | 0 | 13 |
| | male (61–88) | 91 | 2.39 | 2.93 | 0.31 | 0 | 12 |
| | all (21–88) | 444 | 3.00 | 3.28 | 0.16 | 0 | 13 |
| **MADRS** | female (21–40) | 88 | 2.01 | 2.72 | 0.29 | 0 | 12 |
| | male (21–40) | 65 | 2.56 | 2.67 | 0.33 | 0 | 11 |
| | female (41–60) | 72 | 2.27 | 2.70 | 0.32 | 0 | 10 |
| | male (41–60) | 34 | 2.12 | 2.58 | 0.44 | 0 | 10 |
| | female (61–88) | 94 | 2.24 | 2.35 | 0.24 | 0 | 11 |
| | male (61–88) | 91 | 1.69 | 2.53 | 0.27 | 0 | 12 |
| | all (21–88) | 444 | 2.13 | 2.59 | 0.12 | 0 | 12 |

Listed are the mean values and standard deviations (SD) as well as the standard error of the mean (SEM) and the minimum and the maximum of the descriptive information of the cohort. Values are given for six groups of participants comprising young (aged between 21 and 40 years), middle-aged (aged between 41 and 60 years) and older women and men (aged between 61 and 88 years) as well as the whole group. n denotes the number of participants in each group. EHI = Edinburgh Handedness Inventory (Oldfield, 1971), BDI-II = Beck Depression Inventory, 2nd Edition, MADRS = Montgomery–Åsberg Depression Rating Scale

**Table S2.**

**Descriptive statistics of the performance in the four motor tests (n = 444).**

| | Group [sex (age)] | n | mean | SD | SEM | min. | max. |
|---|---|---|---|---|---|---|---|
| **Purdue Pegboard Test** (dominant hand) | female (21–40) | 88 | 16.10 pins | 1.62 | 0.17 | 12.67 pins | 19.30 pins |
| | male (21–40) | 65 | 15.03 pins | 1.63 | 0.21 | 12.00 pins | 20.00 pins |
| | female (41–60) | 72 | 14.88 pins | 1.48 | 0.17 | 11.33 pins | 18.00 pins |
| | male (41–60) | 34 | 14.13 pins | 1.36 | 0.23 | 11.00 pins | 16.67 pins |
| | female (61–88) | 94 | 13.05 pins | 2.01 | 0.21 | 6.24 pins | 18.50 pins |
| | male (61–88) | 91 | 11.96 pins | 1.94 | 0.20 | 7.00 pins | 17.33 pins |
| **Purdue Pegboard Test** (non-dominant hand) | female (21–40) | 88 | 14.95 pins | 1.58 | 0.17 | 14.95 pins | 18.70 pins |
| | male (21–40) | 65 | 14.06 pins | 1.55 | 0.19 | 11.33 pins | 18.33 pins |
| | female (41–60) | 72 | 13.83 pins | 1.60 | 0.19 | 10.33 pins | 18.00 pins |
| | male (41–60) | 34 | 13.33 pins | 1.62 | 0.28 | 10.00 pins | 16.67 pins |
| | female (61–88) | 94 | 12.30 pins | 2.18 | 0.23 | 5.33 pins | 19.00 pins |
| | male (61–88) | 91 | 11.48 pins | 1.97 | 0.21 | 6.67 pins | 16.67 pins |
| **Jebsen-Taylor Test** (dominant hand) | female (21–40) | 88 | 24.71 s | 2.71 | 0.29 | 18.90 s | 33.70 s |
| | male (21–40) | 65 | 26.41 s | 3.56 | 0.44 | 19.82 s | 34.43 s |
| | female (41–60) | 72 | 26.44 s | 4.24 | 0.50 | 19.76 s | 44.5 s |
| | male (41–60) | 34 | 26.54 s | 3.36 | 0.58 | 19.23 s | 33.47 s |
| | female (61–88) | 94 | 29.92 s | 4.71 | 0.49 | 22.45 s | 48.68 s |
| | male (61–88) | 91 | 30.86 s | 5.02 | 0.53 | 21.06 s | 41.21 s |
| **Jebsen-Taylor Test** (non-dominant hand) | female (21–40) | 88 | 26.55 s | 3.55 | 0.38 | 20.67 s | 38.50 s |
| | male (21–40) | 65 | 28.05 s | 3.84 | 0.48 | 20.62 s | 39.90 s |
| | female (41–60) | 72 | 27.74 s | 4.72 | 0.56 | 19.67 s | 51.79 s |
| | male (41–60) | 34 | 27.84 s | 4.07 | 0.70 | 20.82 s | 38.09 s |
| | female (61–88) | 94 | 31.93 s | 5.47 | 0.56 | 22.77 s | 58.69 s |
| | male (61–88) | 91 | 32.42 s | 5.51 | 0.58 | 22.41 s | 52.19 s |

| | | | | | | | |
|---|---|---|---|---|---|---|---|
| **Grip strength (dominant hand)** | female (21–40) | 88 | 58.87 kPa | 11.17 | 1.19 | 28.00 kPa | 89.00 kPa |
| | male (21–40) | 65 | 83.01 kPa | 15.93 | 1.98 | 50.00 kPa | 134.70 kPa |
| | female (41–60) | 72 | 53.01 kPa | 10.78 | 1.27 | 16.67 kPa | 73.00 kPa |
| | male (41–60) | 34 | 72.27 kPa | 14.12 | 2.42 | 47.33 kPa | 95.33 kPa |
| | female (61–88) | 94 | 47.05 kPa | 9.65 | 1.00 | 15.00 kPa | 68.00 kPa |
| | male (61–88) | 91 | 66.19 kPa | 13.39 | 1.40 | 32.00 kPa | 94.00 kPa |
| **Grip strength (non-dominant hand)** | female (21–40) | 88 | 55.22 kPa | 12.01 | 1.28 | 20.00 kPa | 86.67 kPa |
| | male (21–40) | 65 | 79.31 kPa | 17.25 | 2.14 | 39.33 kPa | 129.30 kPa |
| | female (41–60) | 72 | 50.60 kPa | 10.99 | 1.30 | 17.00 kPa | 74.67 kPa |
| | male (41–60) | 34 | 70.27 kPa | 15.22 | 2.61 | 34.67 kPa | 98.67 kPa |
| | female (61–88) | 94 | 44.45 kPa | 9.48 | 0.98 | 17.33 kPa | 68.00 kPa |
| | male (61–88) | 91 | 63.63 kPa | 13.39 | 1.56 | 28.67 kPa | 116.67 kPa |
| **Finger-tapping frequency (dominant hand)** | female (21–40) | 88 | 6.51 Hz | 0.98 | 0.10 | 4.20 Hz | 8.89 Hz |
| | male (21–40) | 65 | 6.65 Hz | 1.17 | 0.15 | 3.96 Hz | 9.54 Hz |
| | female (41–60) | 72 | 6.11 Hz | 1.01 | 0.12 | 2.65 Hz | 8.44 Hz |
| | male (41–60) | 34 | 6.72 Hz | 0.99 | 0.17 | 4.46 Hz | 9.53 Hz |
| | female (61–88) | 94 | 5.44 Hz | 0.96 | 0.10 | 2.57 Hz | 7.57 Hz |
| | male (61–88) | 91 | 5.68 Hz | 1.13 | 0.12 | 2.17 Hz | 7.54 Hz |
| **Finger-tapping frequency (non-dominant hand)** | female (21–40) | 88 | 6.10 Hz | 0.88 | 0.09 | 3.89 Hz | 8.36 Hz |
| | male (21–40) | 65 | 6.32 Hz | 1.10 | 0.14 | 4.24 Hz | 9.44 Hz |
| | female (41–60) | 72 | 5.58 Hz | 0.87 | 0.10 | 3.79 Hz | 7.27 Hz |
| | male (41–60) | 34 | 6.03 Hz | 1.09 | 0.19 | 3.90 Hz | 8.94 Hz |
| | female (61–88) | 94 | 4.96 Hz | 0.79 | 0.08 | 2.77 Hz | 7.04 Hz |
| | male (61–88) | 91 | 5.39 Hz | 1.00 | 0.11 | 2.59 Hz | 7.44 Hz |

Listed are the mean values and standard deviations (SD) as well as the standard error of the mean (SEM) and the minimum and the maximum for the performance in a given motor test with either the dominant or non-dominant hands. Values are given for six groups of participants comprising young (aged between 21 and 40 years), middle-aged (aged between 41 and 60 years) and older women and men (aged between 61 and 88 years). N denotes the number of participants in each group.

**Table S3:**

**Principal component analysis (PCA) revealed three robust motor components for young, middle-aged, and older adults (n = 444).**

| | **Component 1 (*dexterity*)** | | | **Component 2 (*force*)** | | | **Component 3 (*speed*)** | | |
|---|---|---|---|---|---|---|---|---|---|
| | 21-40 | 41-60 | 61-88 | 21-40 | 41-60 | 61-88 | 21-40 | 41-60 | 61-88 |
| Purdue Pegboard Test (dominant) | **0.83** | **0.79** | **0.88** | -0.17 | -0.08 | -0.03 | 0.08 | 0.03 | -0.11 |
| Purdue Pegboard Test (non-dominant) | **0.75** | **0.79** | **0.90** | 0.12 | 0.04 | 0.01 | 0.01 | 0.15 | -0.06 |
| Jebsen-Taylor Test (dominant) | **-0.81** | **-0.81** | **-0.86** | -0.01 | -0.17 | -0.09 | -0.10 | -0.23 | -0.14 |
| Jebsen-Taylor Test (non-dominant) | **-0.79** | **-0.81** | **-0.82** | -0.04 | -0.23 | -0.13 | -0.28 | -0.11 | -0.24 |
| Grip strength (dominant) | -0.11 | 0.07 | 0.07 | **0.98** | **0.96** | **0.98** | 0.01 | 0.12 | 0.05 |
| Grip strength (non-dominant) | -0.06 | 0.06 | 0.05 | **0.98** | **0.97** | **0.97** | 0.01 | 0.11 | 0.09 |
| Finger-tapping (dominant) | 0.15 | 0.24 | 0.03 | -0.02 | 0.11 | -0.01 | **0.95** | **0.87** | **0.94** |
| Finger-tapping (non-dominant) | 0.14 | 0.07 | 0.07 | 0.04 | 0.12 | 0.16 | **0.94** | **0.91** | **0.93** |
| Explained variance (after rotation) in % | 38.0 (32.3) | 40.5 (32.8) | 39.6 (37.6) | 24.7 (24.5) | 23.0 (24.8) | 25.7 (24.3) | 17.7 (23.6) | 15.4 (21.3) | 19.5 (23.0) |
| Eigenvalues (after rotation) | 3.04 (2.58) | 3.24 (2.62) | 3.17 (3.00) | 1.98 (1.96) | 1.84 (1.99) | 2.06 (1.94) | 1.41 (1.89) | 1.24 (1.71) | 1.56 (1.84) |

The principal component analysis (PCA; rotated component matrix) revealed the relationship between the performances of the 153 young (21 years – 40 years), 106 middle-aged (41 years – 60 years), and 185 older participants (61 years – 88 years) in 4 motor tests performed with their dominant and non-dominant hand.

The three components accounted together for 80.4% (young), 79.0% (middle-aged) and 84.8% (older) of the total variances.

Component loadings (after varimax rotation) above 0.7 are highlighted and printed in bold, with the corresponding (sub)tests considered relevant for that component.

**Table S4:**

**Principal component analysis (PCA) revealed three robust motor components for both sexes (n = 444).**

| | Component 1 (*dexterity*) | | Component 2 (*force*) | | Component 3 (*speed*) | |
|---|---|---|---|---|---|---|
| | w | m | w | m | w | m |
| Purdue Pegboard Test (dominant) | **0.83** | **0.85** | 0.17 | 0.23 | 0.22 | 0.09 |
| Purdue Pegboard Test (non-dominant) | **0.86** | **0.89** | 0.20 | 0.14 | 0.16 | 0.11 |
| Jebsen-Taylor Test (dominant) | **-0.85** | **-0.85** | -0.18 | -0.15 | -0.21 | -0.20 |
| Jebsen-Taylor Test (non-dominant) | **-0.84** | **-0.79** | -0.15 | -0.17 | -0.21 | -0.30 |
| Grip strength (dominant) | 0.24 | 0.22 | **0.94** | **0.95** | 0.07 | 0.09 |
| Grip strength (non-dominant) | 0.19 | 0.21 | **0.95** | **0.95** | 0.07 | 0.09 |
| Finger-tapping (dominant) | 0.23 | 0.20 | 0.06 | 0.02 | **0.92** | **0.93** |
| Finger-tapping (non-dominant) | 0.25 | 0.22 | 0.08 | 0.14 | **0.91** | **0.93** |
| Explained variance (after rotation) in % | 53.3 (38.4) | 51.3 (37.5) | 18.1 (24.0) | 18.7 (24.4) | 13.3 (23.0) | 15.4 (23.4) |
| Eigenvalues (after rotation) | 4.23 (3.07) | 4.12 (3.00) | 1.50 (1.92) | 1.49 (1.95) | 1.06 (1.84) | 1.12 (1.88) |

The principal component analysis (PCA; rotated component matrix) revealed the relationship between the performances of the 254 women and 190 men in four motor tests performed with their dominant and non-dominant hands.

The three components accounted for 84.7% (women) and 85.4 (men) of the total variance.

Component loadings (after varimax rotation) above 0.7 are highlighted and printed in bold, with the corresponding (sub)tests considered relevant for that component.
